# Determinants of self-treatment with antimalarials in Ndola district, Zambia: a cross-sectional study

**DOI:** 10.1101/2024.09.02.24312958

**Authors:** Namasiku Grace Susiku, Choolwe Jacobs, Jessy Zgambo, Patrick Kaonga, Lungowe Sitali

**Affiliations:** Department of Epidemiology and Biostatistics, School of Public Health, University of Zambia, Lusaka, Zambia; Department of Biomedical Sciences, School of Health Sciences, University of Zambia, Lusaka, Zambia

**Keywords:** self-medication, drug resistance, prevalence, factors

## Abstract

Antimalarial drug resistance has been an obstacle in the fight against malaria over the years. Antimalarial self-medication is one of the factors associated with antimalarial resistance, and is on the rise globally and has become quite common among developing populations. Self-medication is when individuals choose and use medications to treat symptoms they perceive or diagnose themselves, without the guidance of a healthcare professional. This study aimed at investigating prevalence and determinants of antimalarial drug self-medication among adults in Ndola district, Zambia. A cross-sectional study was conducted among adults in Ndola district, Zambia in June 2023. Using multistage cluster sampling, 446 participants were randomly selected and interviewed through a mobile-based structured questionnaire administered by the researchers using Kobo collect/kobo toolbox. The head of household was the primary participant. Data was then exported to Microsoft Excel, cleaned and coded, and thereafter exported to STATA version 16.1 for analysis. Chi-square tests and logistic regression analysis was used to test associations between categorical variables and to estimate the odds of antimalarial drug self-medication associated with the explanatory variables. The prevalence of antimalarial drug self-medication was 20% in this study population and the factors significantly associated were; moderate to mild illness, bad experience with hospital care, lack of knowledge about antimalarial drug resistance, and the perception that antimalarial drug self-medication was not risky. The finding that antimalarial self-medication was common in this study population is suggestive of a poor state of the health-care delivery system possibly due to inadequate manpower, stock out of drugs in health facilities, and lack of awareness about the risks of self-medication. This further suggests that there is great need for regulatory authorities to strengthen drug regulations and update the list of over-the-counter and essential medicines to ensure the safety of public health. In addition, authorities should repackage malaria sensitization messages to strongly highlight the risks of antimalarial drug self-medication.

## Introduction

Malaria is a preventable disease that is not so costly to treat, yet continues to thrive as an issue of public health concern. As of the year 2021, the World Health Organisation (WHO) estimated about 247 million malaria cases globally, an increase from 245 million in 2020 with most of this increase coming from countries in the WHO African region. The malaria mortality rate was 15.1 deaths per 100,000 of the population at risk in 2020 before decreasing slightly to 14.8 in 2021 (1). Up to 35.4 million disability-adjusted life years (DALYs) are lost in the sub-Saharan Africa region alone due to malaria mortality and morbidity (2). Zambia carries 2% of the global malaria case burden and in 2015, an estimated 4 million confirmed malaria cases and 2,389 reported deaths were due to malaria in Zambia alone (3). Interventions that have been employed to reduce the burden of malaria include; the scale-up of vector control interventions, including long-lasting insecticidal nets (LLINs), and indoor residual spraying of insecticide (IRS), coupled with improved case management using artemisinin-based combination therapy (ACT), and intermittent preventive therapy with Sulfadoxine-pyrimethamine (SP) for pregnant women (4).

However, antimalarial drugs are prone to self-medication in settings where they are not well-regulated and have easy access to over-the-counter medications without prescription. Antimalarial drug self-medication can be defined as taking anti-malarial drugs on an individual’s initiative without consulting a qualified prescriber. Several advantages have been associated with appropriate self-medication, including enhanced access to medication and relief for patients, increased patient involvement in their healthcare, more effective utilization of healthcare professionals’ expertise, and reduced or optimized governmental healthcare expenditure related to the treatment of minor health issues. However, self-medication is not without its risks, particularly in cases of irresponsible self-medication. Antimalarial drug self-medication can lead to parasite resistance to antimalarial drugs that has been reported (3) in many malaria-endemic countries. Additionally, it can lead to delays in seeking necessary medical advice and masking of serious illnesses, resulting in paradoxical economic loss due to late diagnosis of malaria with adverse outcomes. Antimalarial drug resistance has substantially limited treatment options and is now the greatest obstacle to controlling the disease worldwide (5).

Previous studies have demonstrated that antimalarial drug self-medication is common among developing populations from as low as 4.4% to as high as 80% (6–10). Some factors associated with antimalarial drug self-medication are age, female gender, marital status, far distance to the health facility, lack of money, higher level of education, experience with similar disease, bureaucracies at the health facility, lack of knowledge, mild illness, experience with treating similar disease, risk perception, easy access to antimalarial drugs and health insurance among others (6, 7, 11–15).

Although some studies have demonstrated the prevalence of self-medication and associated factors in Zambia (16–19), there are limited studies regarding the prevalence and factors associated with antimalarial drug self-medication at community level in Ndola district, Zambia. Therefore, this study aimed at investigating prevalence of antimalarial drug self-medication and associated factors among adults in Ndola district, Zambia.

## Methods

### Study design and setting

A cross-sectional study design was used to determine the prevalence and factors associated with antimalarial drug self-medication among adults in Ndola district, Zambia between November 2022, and March 2023. The study was conducted in Ndola district located in Copperbelt province, Zambia as shown in figure 1. The prevalence of malaria in Copperbelt province ranges from 8% to 11%. Ndola district has a total area of 965.3 km² and as of the 2010 census, its total population was estimated to be 451,246 (223,020 males and 228,226 females), with a population density of 467.5/km² (Census, 2010). The district is politically divided into 28 wards namely, Chichele, Chifubu, Chipulukusu, Daj Hammarskjold, Fibobe, Itawa, Kabushi, Kafubu, Kaloko, Kamba, Kaniki, Kanini, Kansenshi, Kantolomba, Kavu, Kawama, Lubuto, Masala, Mukuba, Munkulungwe, Mushili, Nkwazi, Pamodzi, Skyways, Toka, Twapia, Twashuka and Yengwe. Only eight of these wards were included in the study by random selection and these were Lubuto, Itawa, Chifubu, Chipulukusu, kansenshi, Nkwazi, Kafubu, and Twapia. The selected wards accounted for 29% of all the wards thus, were representative of the total population.

**Figure 1.**
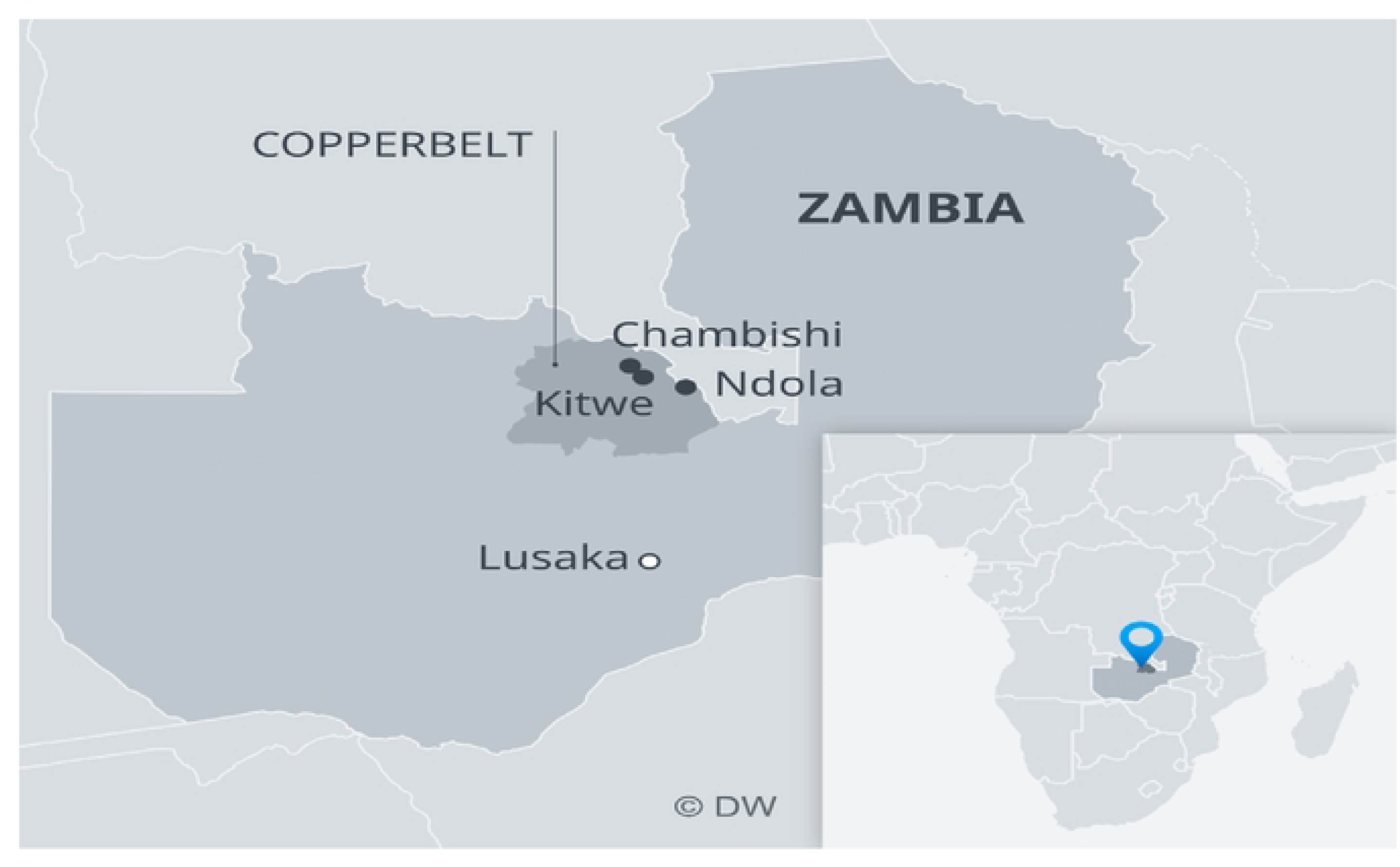

### Study population and sampling strategy

The study population was adult residents of Ndola district, Copperbelt province, Zambia. Since the study population was spread across a wide geographical area, a multistage cluster sampling was used. The sample size was calculated using the proportion formula which gave a minimum sample size of 372 to which 20% was added to account for non-response which brought the total sample size to 447. Probability proportionate to size sampling was used to allocate a representative sample to each of the wards. Thereafter, systematic random sampling was employed to select the households from which participants were sampled in each ward. A central location was selected in each ward at which a direction to sample from was randomly selected. The lottery method was used to select the first house from which a participant was randomly selected and the calculated Kth interval was used to skip households in the selected direction of the central point. At every systematically selected household, we requested for the head of household to interview, in whose absence another participant was then selected. Where the selected participant did not meet the eligibility criteria, another participant from the same household who was eligible to participate in the study was randomly selected. This was done in all selected households in each ward until the sample size was met.

### Data collection tools and procedures

Data was collected primarily through structured interviews using a pre-tested structured questionnaire. Prior to administering the questionnaire, the interviewer presented different antimalarial drugs to the participants to find out if they had used any antimalarials between November 2022 and March 2023. This was done to fulfill the eligibility criteria and prevent a waste of resources. Antimalarial drug Self-medication practice, the outcome variable, was assessed by the selection and use of antimalarial drugs by individuals or a member of the individual’s family without a physician’s order to treat self-recognized or self-diagnosed conditions in the past 6 months. Data collection was also done during evening hours to capture those who worked during the day.

### Description of variables

#### Dependent/outcome Variable

The outcome variable was “antimalarial drug self-medication” which was dichotomized as “yes” if the respondent did not seek medical advice before antimalarial medication and “no” if the respondent sought medical advice before antimalarial medication.

#### Independent/ explanatory variables

These include age, gender, marital status, level of education, employment status, monthly income, severity of illness, underlying health conditions, duration of symptoms prior to self-medication, name of drug taken, health insurance, ready access to health facility, ready access to drugs, past experience with hospital care, level of knowledge about antimalarial drug resistance and risk perception around antimalarial drug self-medication.

### Data management and analysis

To increase efficiency, reduce cost, and ensure quality of the data, kobo toolbox/Kobo collect was used to administer the structured questionnaire (20). The mobile-based data collection method was ideal for enhancing credibility of the data as it incorporates various quality-checking tools including completeness and consistency checks (20). Thereafter, Data was directly exported to STATA version 16.0 for analysis. Microsoft Excel spreadsheets containing variables of interest were imported into STATA version 16.1 MP (Stata Corporation, College Station, Texas, USA) for analysis. At analysis, descriptive statistics using frequencies were reported for Categorical variables. The Pearson’s Chi-square test was used to investigate association between categorical variables. A mixed-effect logistic regression was used to perform multivariable analysis to estimate the odds of antimalarial drug self-medication associated with the explanatory variables. A mixed-effect model was preferred for this particular study to account for clustering effect so that the results can be used to make inferences on the entire district and not just on the study participants (21). An investigator-led stepwise approach was undertaken to fit the best model using the likelihood ratio tests coupled with the Akaike Information Criterion (AIC) and the Bayesian Information Criterion (BIC). Multicollinearity of the models was checked using the tolerance and inflation statistics to fulfill the law of parsimony. Sensitivity and specificity tests were run to check the performance of the best model and to ascertain that the model was not classifying participants by chance, the Receiver Operations Characteristic Curve (ROC) was used.

### Ethical consideration

Ethical clearance was sought and granted from the University of Zambia Biomedical Research Ethics Committee (UNZABREC) REF. No. 3603-2023, and the National Health Research Authority (NHRA) Ref No. NHRA015/03/07/2023. Permission to conduct the study was also sought and granted from Ndola District Health Office. Detailed study information sheets were availed to the participants prior to obtaining informed consent. Participants were allowed to withdraw from the study at any point during the interview and no individual was coerced to participate in the study. Permission was obtained from the head of the household even when he/she would not be the primary respondent. The study maintained the highest level of confidentiality as participants were identified by identification numbers rather than their names nor other personal information.

## Results

### Social demographic characteristics of the adult participants of Ndola district, Zambia; June 2023

Participants’ social demographic characteristics are presented in Table 1. The study included 446 eligible participants aged between 18 and 84 years with the mean age of 37 (± 14) years. About three-quarters (74.2%, 331/446) of the participants were females. Majority (61%, 272/446) were married and 46% (205/446) had attended up to secondary education. Above two-thirds (69.7%, 311/446) were unemployed. About 70.2% (313/446) of the participants did not earn a monthly income.

**Table 1:**
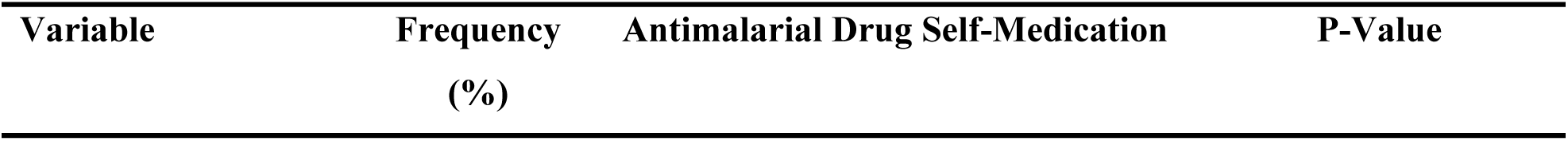
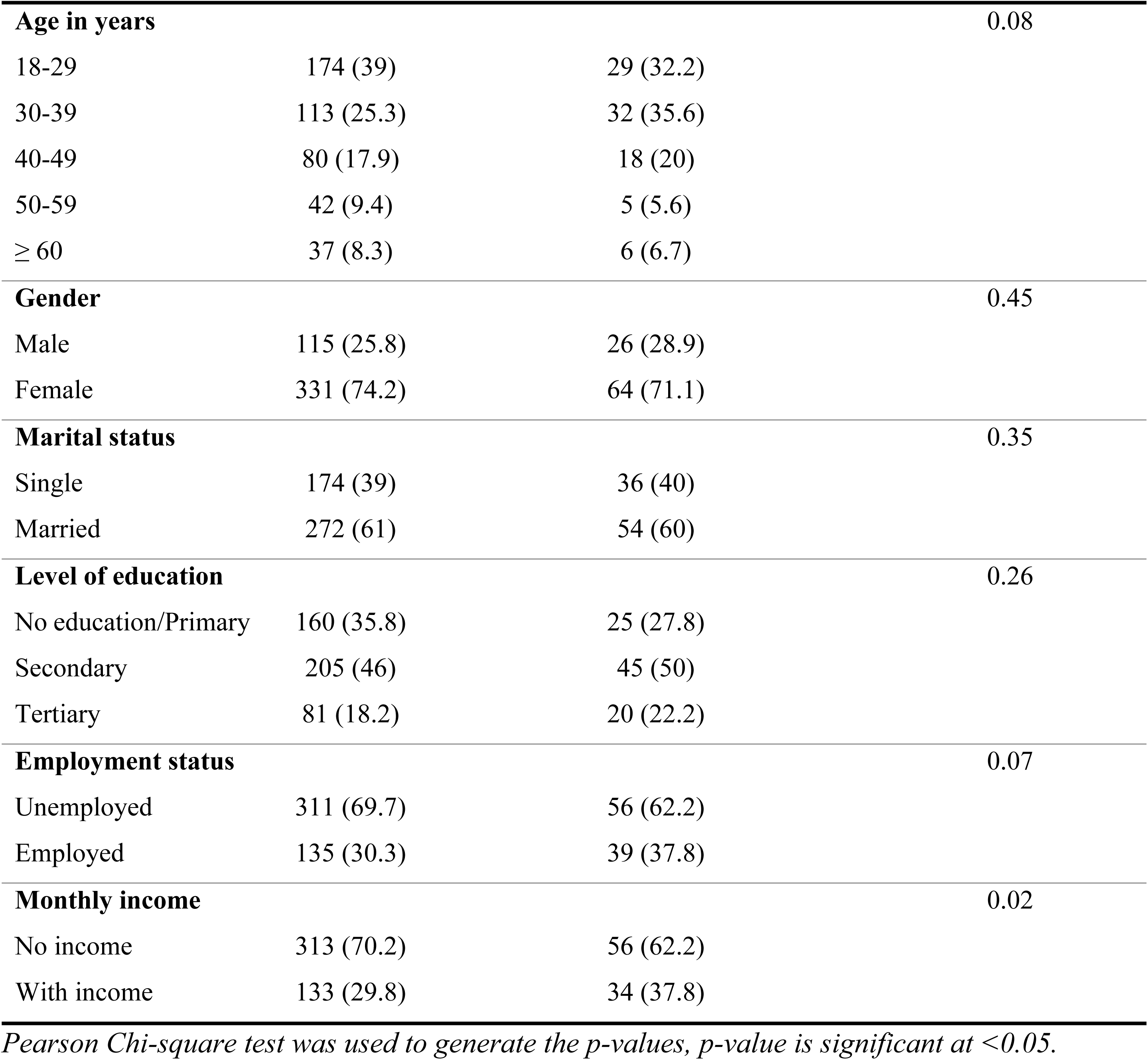
Social demographic characteristics of adult participants of Ndola District, Zambia; June 2023.

| Variable | Frequency<br>(%) | Antimalarial Drug Self-Medication | P-Value |
| --- | --- | --- | --- |
| **Age in years** |  |  | 0.08 |
| 18-29 | 174 (39) | 29 (32.2) |  |
| 30-39 | 113 (25.3) | 32 (35.6) |  |
| 40-49 | 80 (17.9) | 18 (20) |  |
| 50-59 | 42 (9.4) | 5 (5.6) |  |
| ≥ 60 | 37 (8.3) | 6 (6.7) |  |
| **Gender** |  |  | 0.45 |
| Male | 115 (25.8) | 26 (28.9) |  |
| Female | 331 (74.2) | 64 (71.1) |  |
| **Marital status** |  |  | 0.35 |
| Single | 174 (39) | 36 (40) |  |
| Married | 272 (61) | 54 (60) |  |
| **Level of education** |  |  | 0.26 |
| No education/Primary | 160 (35.8) | 25 (27.8) |  |
| Secondary | 205 (46) | 45 (50) |  |
| Tertiary | 81 (18.2) | 20 (22.2) |  |
| **Employment status** |  |  | 0.07 |
| Unemployed | 311 (69.7) | 56 (62.2) |  |
| Employed | 135 (30.3) | 39 (37.8) |  |
| **Monthly income** |  |  | 0.02 |
| No income | 313 (70.2) | 56 (62.2) |  |
| With income | 133 (29.8) | 34 (37.8) |  |

*Pearson Chi-square test was used to generate the p-values, p-value is significant at <0.05.*

### Clinical characteristics of adult participants of Ndola district, Zambia; June 2023

Participants’ clinical characteristics are displayed in Table 2. Among the 446 participants, 189 (42.2%) reported having moderate to mild illness before medication. The majority, 366/446 (82.1%) of the participants did not suffer from any underlying health condition. The larger proportion 359/446 (80.8%), of participants in this study experienced malaria symptoms for more than twenty-four hours before medication and more than three quarters, 383/446 (85.9%) of the participants took ACT. Three Hundred and Eighty-six (86.6%, 386/446)) of the participants perceived antimalarial drug self-medication as being risky.

**Table 2:**
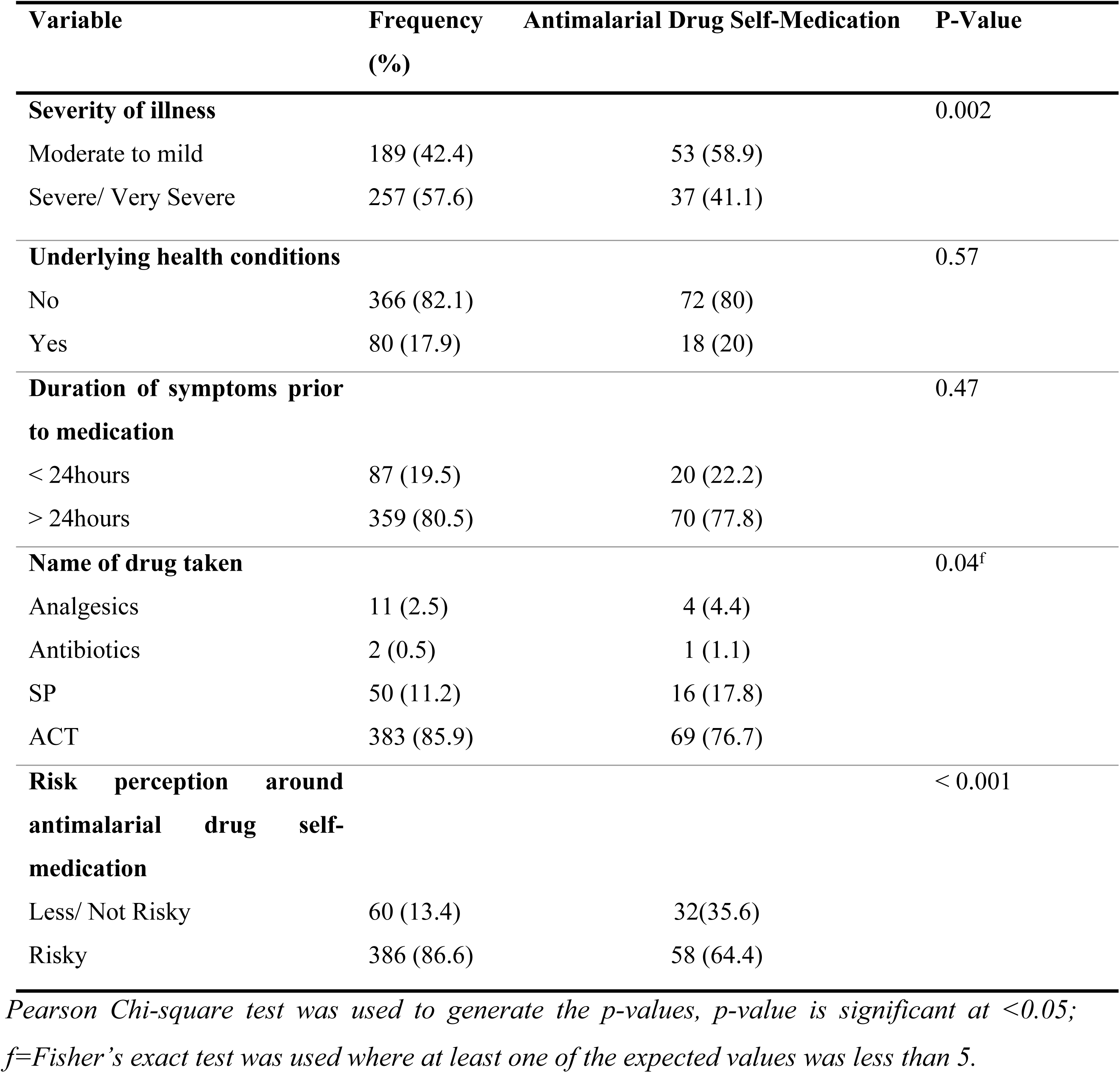
Clinical characteristics of adult participants of Ndola District, Zambia; June 2023.

### Healthcare-related characteristics of adult participants of Ndola district, Zambia; June 2023

Healthcare-related characteristics of the study participants are presented in Table 3. The majority of the participants had ready access to drugs and health facilities, 408/446 (91.5%) and 393/446 (88.1%) respectively. The larger proportion, 343/446 (76.9%) of participants were not on health insurance. Most, 388/446 (87%), of the participants in this study recorded having had good/very good experience with hospital care.

**Table 3:** Healthcare-related characteristics of adult participants of Ndola district, Zambia; June 2023.

| Variable | Frequency (%) | Antimalarial Drug Self-Medication | P-Value |
| --- | --- | --- | --- |
| **Ready access to drugs** |  |  | 0.26 |
| No (> 5km away) | 38 (8.5) | 5 (5.6) |  |
| Yes (< 5km away) | 408 (91.5) | 85 (94.4) |  |
| **Ready access to health facility** |  |  | 0.33 |
| No (> 5km away) | 53 (11.9) | 8 (8.9) |  |
| Yes (< 5km away) | 393 (88.1) | 82 (91.1) |  |
| **Health insurance** |  |  | 1 |
| No | 343 (76.9) | 69 (67.7) |  |
| Yes | 103 (23.1) | 21 (32.3) |  |
| **Past experience with hospital care** |  |  | < 0.001 |
| Bad/Very Bad | 58 (13) | 25 (27.8) |  |
| Good/ Very Good | 388 (87) | 65 (72.2) |  |

Pearson Chi-square test was used to generate the p-values, p-value is significant at <0.05.

### Prevalence of antimalarial drug self-medication

Out of the 446 participants in this study, about 90 self-medicated with antimalarials (20%; 95% CI 0.17-0.24). Antimalarial drug self-medication was most common among participants who were; female, aged between 30 and 39 years old, married, unemployed, attended secondary education, suffered moderate to mild illness prior to medication, did not have underlying health conditions, experienced symptoms for more than twenty four hours before medication, took ACTs, perceived antimalarial drug self-medication as being not risky, had ready access to drugs and health facilities and those that had good experience with hospital care.

### Factors associated with antimalarial drug self-medication

Results from the univariate and multivariable logistic regression are presented in Table 4. The association between severity of illness and antimalarial drug self-medication was that, suffering from severe illness significantly reduced the chance of antimalarial drug self-medication by 52% compared to suffering from moderate to mild illness (AOR=0.48, 95% CI 0.29-0.79, p-value = 0.004). Having good experience with hospital care significantly reduced the odds of antimalarial drug self-medication by a factor of 0.28 compared to having bad experience (AOR=0.28, 95% CI 0.15-0.52, p-value ˂ 0.001). Additionally, the risk of antimalarial drug self-medication among participants who were knowledgeable about antimalarial drug resistance was 0.48 times lower than the risk of antimalarial drug self-medication among participants who were not knowledgeable (AOR=0.48, 95% CI 0.28-0.8, p-value = 0.005). Finally, the odds of antimalarial drug self-medication among participants who perceived antimalarial drug self-medication to be risky reduced by 84% compared to those who perceived antimalarial drug self-medication as not being risky (AOR=0.26, 95% CI 0.12-0.58, p-value = 0.001).

**Table 4:** Factors Associated with Antimalarial Drug Self-medication Among Adults in Ndola District, Zambia.

| Variable | Crude Estimates |  | Adjusted Estimates |  |
| --- | --- | --- | --- | --- |
|  | OR (95% CI) | p-value | AOR (95% CI) | p-value |
| **Monthly income** |  |  |  |  |
| No income | Ref | N/A | Ref | N/A |
| With income | 1.31 (1.08-1.6) | 0.007 | 1.95 (1.19-3.2) | 0.008 |
| **Severity of illness** |  |  |  |  |
| Moderate to mild | Ref | N/A | Ref | N/A |
| Severe | 0.43 (0.27-0.69) | <0.001 | 0.48 (0.29-0.79) | **0.004** |
| **Ready access to health facility** |  |  |  |  |
| No (> 5km away) | Ref | N/A | Ref | N/A |
| Yes (< 5km away) | 1.48 (0.67-3.27) | 0.33 | 2.22 (0.73-6.75) | 0.16 |
| **Ready access to drugs** |  |  |  |  |
| No (> 5km away) | Ref | N/A | Ref | N/A |
| Yes (< 5km away) | 1.74 (0.66-4.58) | 0.27 | 0.86 (0.24-3.11) | 0.82 |
| **Past experience with hospital care** |  |  |  |  |
| Bad | Ref | N/A | Ref | N/A |
| Good | 0.27 (0.15-0.48) | <0.001 | 0.28 (0.15-0.52) | **<0.001** |
| **Knowledge about antimalarial drug resistance** |  |  |  |  |
| Not knowledgeable | Ref | N/A | Ref | N/A |
| Knowledgeable | 0.39 (0.24-0.63) | < 0.001 | 0.48 (0.28-0.8) | **0.005** |
| **Risk perception around antimalarial drug self-medication** |  |  |  |  |
| Not /Less Risky | Ref | N/A | Ref | N/A |
| Risky | 0.19 (0.57-2.27) | <0.001 | 0.26 (0.12-0.58) | **0.001** |

*Data presented as odds ratios (OR) with confidence intervals (CI); p-value is significant at < 0.05,*
*Ref: Reference group (category), AOR; Adjusted Odds Ratio.*

## Discussion

The prevalence of antimalarial drug self-medication was common at 20% and could be as low as 17% and as high as 24% among adults in Ndola district, Zambia. Factors that were statistically significantly associated with antimalarial drug self-medication are; moderate to mild illness, bad experience with hospital care, lack of knowledge about antimalarial drug resistance, and the perception that antimalarial drug self-medication is not risky.

The finding that self-medication with antimalarial drugs is common (20%; 95% CI 0.17-0.24) in our study population is in line with what other studies found in similar settings; 22.6% in Nigeria (22) and 19% in Tanzania (23). Contrary to this, some studies found a higher prevalence such as 43.4% among community members in Khartoum state, Sudan (11), 79.5% among undergraduate students in Khartoum state, Sudan (7) and 49.3% in Cameroon (6), probably due to differences in methodologies and study populations. This finding suggests a poor healthcare delivery system in Ndola district, Zambia. A shortage of consumables in health facilities could be a reason for the high prevalence of antimalarial drug self-medication. People visit health facilities with the hope that they will return home with a solution to their illness such as medication. However, in circumstances where there is a stock out of drugs in health facilities, people tend to shun away from visiting health facilities for medical advice when unwell, rather they purchase medication from pharmacies and other sources because they want to save time. Long queuing time at health facilities is another reason for antimalarial drug self-medication, cited by participants in this study. Furthermore, participants in this study who had suffered from malaria in the past expressed a high level of certainty about malaria symptoms and would claim to have malaria whenever they experienced such symptoms, prompting them to self-medicate. The prevalence of antimalarial drug self-medication realized from this study further suggests that there is an urgent need for regulatory authorities to strengthen drug regulations and update the list of over-the-counter and essential medicines to ensure the safety of consumers.

The finding that participants who suffered from moderate to mild illness were more likely to self-medicate with antimalarial drugs is similar to what was found in Cameroon (6) and China (24) despite the difference in study populations. A possible explanation for this could be that when the symptoms are mild, people opt to use their experience with treatment of similar symptoms to self-medicate rather than visiting the health facility for a diagnosis. However, when the disease is severe, the amount of pain felt prompts the patient to visit a health facility for treatment for the fear of dying in their home. This study also revealed that participants who had very bad experience with hospital care were significantly more likely to self-medicate with antimalarial drugs, similar to what was found in China (24). This finding reflects inefficiency in the primary delivery of healthcare possibly due to inadequate manpower and poor work culture of health professionals. It is quite sensible that when a person has had a bad experience with hospital care, they would never want to go back there, rather they will opt to self-medicate. This finding suggests that there may be need to scale up man-power in health facilities for efficiency.

In most instances, an individual will not indulge in an act or behaviour which they perceive to be risky (25). Therefore, people that perceive antimalarial drug self-medication to be very risky are less likely to self-medicate due to the fear of adverse health effects as observed in this study. This finding is in line with what a similar study found in Saudi Arabia (26). Finally, a lack of knowledge about antimalarial drug resistance was found to significantly increase the odds of antimalarial drug self-medication in this study population. This is in relation to the fact that self-medication is linked to drug resistance. However, this finding is contrary to what was found in Nepal (27) probably due to the differences in methodology and study populations.

Previous studies found significant associations between antimalarial drug self-medication and factors such as female gender, marital status, lack of income, ready access to drugs, far distance to the health facility and health insurance. On the contrary, none of these associations were significant in this study most likely due to differences in study populations, sample sizes and methodologies.

## Conclusion

Antimalarial drug self-medication is common among adults in Ndola district, Zambia and the factors associated are: moderate to mild illness, bad experience with hospital care, lack of knowledge about antimalarial drug resistance, and the perception that antimalarial drug self-medication is not risky. It is worth noting that, although this study has successfully demonstrated the prevalence of antimalarial drug self-medication and associated factors in Ndola district, one participant was not included in the analysis due to incomplete responses. However, this was accounted for by the large sample size which increased the power of this study, making the results statistically reliable. It would be critical to inform and educate the population on the dangers of self-medication and for regulatory authorities to strengthen drug regulations and update the list of over-the-counter and essential medicines to ensure the safety of citizens. Authorities need also to ensure that there is constant supply of consumables in health facilities such as antimalarial drugs to encourage people to seek medical advice when ill.

## Data Availability

A data set and do file used for analysis is readily available and we have no problems availing it whenever necessary

## Author Contributions

**Conceptualization:** Namasiku Grace Susiku, Lungowe Sitali-Zimba, Choolwe Jacobs, Jessy Zgambo-Akatumwa

**Data curation:** Namasiku Grace Susiku

**Formal analysis:** Namasiku Grace Susiku

**Investigation:** Namasiku Grace Susiku

**Methodology:** Namasiku Grace Susiku, Lungowe Sitali-Zimba, Choolwe Jacobs, Jessy Zgambo-Akatumwa

**Project administration:** Namasiku Grace Susiku

**Resources:** Namasiku Grace Susiku

**Software:** Namasiku Grace Susiku

**Supervision:** Namasiku Grace Susiku, Lungowe Sitali-Zimba, Choolwe Jacobs, Jessy Zgambo-Akatumwa

**Validation:** Namasiku Grace Susiku, Lungowe Sitali-Zimba, Choolwe Jacobs, Jessy Zgambo-Akatumwa

**Visualization:** Namasiku Grace Susiku, Lungowe Sitali-Zimba, Choolwe Jacobs, Jessy Zgambo-Akatumwa

**Writing - original draft:** Namasiku Grace Susiku, Lungowe Sitali-Zimba, Choolwe Jacobs, Jessy Zgambo-Akatumwa, Patrick Kaonga

**Writing – review & editing:** Namasiku Grace Susiku, Lungowe Sitali-Zimba, Choolwe Jacobs, Jessy Zgambo-Akatumwa, Patrick kaonga

## Acknowledgments

Namasiku Grace Susiku is a recipient of the Norwegian Programme for Capacity Development in Higher Education and Research for Development (NORHED PRICE scholarship). We are grateful for the support without whom this study could have not been possible. We further extend our gratitude to the Ndola District Health Office (NDHO) which facilitated the implementation of this study. We are also grateful to all the participants who gave their consent for the study. Finally, we are grateful to members of the faculty of Epidemiology and Biostatistics, University of Zambia, for their guidance throughout this study.

